# High resolution dermal ultrasound (US) combined with superficial radiation therapy (SRT) versus non-image guided SRT or external beam radiotherapy (XRT) in early-stage epithelial cancer: a comparison of studies

**DOI:** 10.1101/2022.08.01.22278255

**Authors:** Lio Yu, Mairead Moloney, Songzhu Zheng, James Rogers

## Abstract

**Background:** To compare the effectiveness of high-resolution dermal ultrasound (US) guided superficial radiotherapy (SRT) to non-image-guided radiotherapy in the treatment of early-stage epithelial cancer.

**Methods:** A high-resolution dermal ultrasound (US) image guided form of superficial radiation therapy (designated here as US-SRT) was developed in 2013 where the tumor configuration and depth can be visualized prior to, during, and subsequent to treatments, using a 22 megahertz (MHz) dermal ultrasound (US) with a doppler component. We previously published the results using this technology to treat 2917 early-stage epithelial cancers showing a high local control (LC) rate of 99.3%. We compared these results with similar American studies from a comprehensive literature search used in an article/guideline published by American Society of Radiation Oncology (ASTRO) on curative radiation treatment of basal cell carcinoma (BCC), squamous cell carcinoma (SCC) and squamous cell carcinoma in-situ (SCCIS) lesions from 1988 to 2018. Only U.S. based studies with greater than 100 cases with similar patient/lesion characteristics and stages treated by external beam, electron, or superficial/orthovoltage radiation therapy were included in the criteria for selection. The resultant 4 studies had appropriate comparable cases identified and the data analyzed/calculated with regard to local control. Logistic regression analysis was performed comparing each study to US-SRT individually and collectively with stratification by histology (BCC, SCC, and SCCIS).

**Results:** US-SRT LC was found to be statistically superior to each of the 4 non-image-guided radiation therapy studies individually and collectively (as well as stratified by histology subtype) with p-values ranging from p< 0.0001 to p= 0.0438.

**Conclusions:** Results of US-SRT in local control were statistically significantly superior across the board versus non-image-guided radiation modalities in treatment of epithelial NMSC and should be considered a new gold standard for treatment of early-stage cutaneous BCC, SCC, and SCCIS.

## Background

Non-melanoma skin cancer (NMSC) is the most common cancer diagnosed in the United States and it is comprised mostly of basal cell carcinoma (BCC), squamous cell carcinoma (SCC), and squamous cell carcinoma in-situ (SCCIS). [1] The most current estimates of NMSC incidence are from 2012, where it was estimated that 5·43 million NMSC lesions in 3·32 million individuals in the U.S. were diagnosed. [2] The incidence is expected to be increasing by two to three percent yearly. [3] In 2022, this translates to 4·05 to 4·66 million individuals in 6·62 to 7·30 million lesions.

Despite NMSC having a low mortality, high cure rates, rare metastasis, and only accounting for 0·1% of cancer deaths, the standard of treatment is surgical removal. [1] Surgical options consist of Mohs micrographic surgery (MMS), standard surgical excision, shave removal, curettage, and electrodessication. [4] However, numerous nonsurgical, non-invasive modalities exist including topical treatments (i.e., imiquimod, 5-flurouracil [5-FU] etc.), cryotherapy, photodynamic therapy (PDT), laser therapy, and radiotherapy with several techniques available within each category. However, surgery, specifically MMS, has remained the mainstay of treatment as the literature promises the highest cure-rates at around 99%. [5] [6]

Multiple radiation modalities exist for the treatment of NMSC, including brachytherapy, electron beam radiotherapy (EBRT), external radiation therapy (XRT), and superficial radiation therapy (SRT). Specifically, superficial radiation therapy, has been used by dermatologists for the past century to treat NMSC. [7] With the advent of MMS, this modality fell out of favor. However, more recently there has been advancements to SRT, including the use of a high frequency 22 megahertz (MHz) dermal ultrasound (US) to visualize superficial depth of the skin. This has led to the development of high-resolution dermal ultrasound image guided superficial radiotherapy, designated here as (US-SRT) and commercially known as image-guided SRT (IGSRT). Commercial units became available in 2014 that allowed for lesion visualization prior to, during, and after treatment. A recently published study by Yu *et al*. using US-SRT (“IGSRT”) to treat 2917 early-stage keratinocytic cancers yielded a high local control (LC) rate of 99·3%. [8] An updated abstract that added 93 patients and 133 lesions for a total of 1725 patients with 3050 early-stage keratinocytic lesions showed a continued high local control (LC) of 99·2%. [9] This suggests that US-SRT can offer comparable cure rates to that of the current “gold standard” treatment modality, namely MMS, for early-stage NMSC without needing surgery and its associated risks, discomfort, and cosmetic sequalae.^A^

## Objective

To statistically evaluate the local control (LC) differences, if any, of US-SRT versus non-image-guided radiotherapy modalities for the treatment of early-stage epithelial cancer.

## Methods

### Source Information

US-SRT results have been investigated in two seminal studies referenced hereafter as the 2021 US-SRT study and the 2022 US-SRT study. [8] [9] Data from the 2021 US-SRT study was obtained via direct chart analysis of patients with histopathologic confirmed NMSC treated with US-SRT from multiple institutions. Data from the 2022 US-SRT study utilized the same data from the 2021 US-SRT study with the addition of 133 histopathological confirmed NMSC from 94 patients with updated follow-up intervals. Data on the additional 133 histopathologic confirmed NMSC in 94 patients was previously published. [10]

US-SRT outcomes included in the present analyses are from a subset of the 2021 US-SRT study patients who have a follow-up of greater than 52 weeks; and are from the entire study population of the 2022 US-SRT study.

The American Society of Radiation Oncology (ASTRO) published a literature review containing 143 studies on curative radiation treatment for NMSC. [11] A subset of modern, pertinent, comparable studies that utilized superficial radiotherapy (SRT) and external radiotherapy (XRT) without image-guidance were identified as meeting the following inclusion/exclusion criteria.

- <u>Inclusion:</u> Only studies performed in the USA
- <u>Exclusion:</u> Meta-analysis, brachytherapy, pre-operative, post-operative +/- chemo/targeted agents used, recurrent or predominately recurrent, prior radiotherapy, T4 only/predominant, metastatic to parotids, involvement of parotids, wound healing only, no local control reported, perineural invasion in ≥50% of cases, and those lesions arising from scar.

These criteria resulted in four high-quality, recent, evidenced-based studies, each with greater than 100 subjects. The four studies provided the XRT/SRT outcome data for this study and are hereafter referenced as the Lovett study, Locke study, Silverman study, and Cognetta study. [12] [13] [14] [15]

### Local Control (LC) Calculation

Lesion counts for the two US-SRT studies and the four XRT/SRT studies were used to compute LC as “Number of lesions that did not recur / Total number of lesions”. Lesions were analyzed as independent events. Only T_is_, T_1_ and T_2_ lesions were included.

### Statistical Analysis

Data availability in the two US-SRT studies and the four XRT/SRT studies made it possible to compare each US-SRT study (i.e., Yu study and Moloney study) to the following XRT/SRT studies: Lovett, Locke, Silverman, and Cognetta for BCC; Lovett, Locke, and Cognetta for SCC; and Cognetta for SCCIS. For each comparison, a logistic model was implemented that contained the effect of treatment with treatment levels defined as the studies under consideration. Odds ratios were then derived that compared the US-SRT studies to each available XRT/SRT study and to all XRT/SRT studies combined.

## Results

Table 1 compares each of the US-SRT studies to each XRT/SRT study (Lovett, Locke, Silverman, and Cognetta).^B^ Comparisons using odds ratios from logistic regression models were made for BCC, SCC, and SCCIS, individually, with odds ratios greater than unity (1) favoring US-SRT. As indicated in Table 1, not all studies evaluated all histologic tumor categories (SCC, SCCIS, BCC), and therefore, only studies that included a given histologic tumor category could be used as a comparator study in the evaluation of that same histologic tumor category. Table 1 also reports comparisons by tumor category between each US-SRT study and all comparator XRT/SRT studies combined. Finally, the LC odds can be converted to a probability with an asymmetrical 95% confidence interval. For the 2021 and 2022 US-SRT studies, separately, LC probabilities and 95% confidence intervals were calculated for each US-SRT study. This allows an odds ratio comparison to be envisioned as a plotted difference in LC probabilities.

**Table 1.** Recurrence and Local Control Events for Basal Cell Carcinoma (BCC), Squamous Cell Carcinoma (SCC) and Squamous Cell Carcinoma In Situ (SCCIS) with High resolution dermal ultrasound image guided superficial radiotherapy (US-SRT) versus External Radiation Therapy (XRT) / Superficial Radiation Therapy (SRT) Contrasts Using Odds Ratios (OR)

| Lesion | Treatment | Study | Outcome |  |  | Odds Ratio<br>(XRT/SRT over US-SRT)<br>(95% Confidence Limits) |  |  |  |
| --- | --- | --- | --- | --- | --- | --- | --- | --- | --- |
|  |  |  | Recurrence |  | Local Control |  |  |  |  |
|  |  |  | (n) | (%) | (n) | Yu 2021: US-SRT |  | Moloney 2022: US-SRT |  |
| BCC | US-SRT | Yu (2021) | 6 | 0.9 | 698 | OR | p-value | OR | p-value |
|  |  | Moloney (2022) | 16 | 1.1 | 1471 |  |  |  |  |
|  | XRT/SRT | Lovett | 20 | 9.0 | 202 | 11.5<br>(4.6, 29.1) | <.0001 | 9.1<br>(4.6, 17.9) | <.0001 |
|  |  | Locke | 21 | 6.4 | 305 | 8.0<br>(3.2, 20.0) | <.0001 | 6.3<br>(3.3, 12.3) | <.0001 |
|  |  | Silverman | 52 | 6.0 | 810 | 7.5<br>(3.2, 17.5) | <.0001 | 5.9<br>(3.3, 10.4) | <.0001 |
|  |  | Cognetta | 22 | 3.1 | 690 | 3.7<br>(1.5, 9.2) | 0.0047 | 2.9<br>(1.5, 5.6) | 0.0012 |
|  |  | All | 115 | 5.4 | 2007 | 7.1<br>(3.1, 16.3) | <.0001 | 5.6<br>(3.3, 9.6) | <.0001 |
| SCC | US-SRT | Yu (2021) | 4 | 0.7 | 544 | OR | p-value | OR | p-value |
|  |  | Moloney (2022) | 7 | 0.8 | 926 |  |  |  |  |
|  | XRT/SRT | Lovett | 14 | 18.9 | 60 | 31.7<br>(10.1, 99.5) | <.0001 | 30.9<br>(12.0, 79.3) | <.0001 |
|  |  | Locke | 15 | 15.2 | 84 | 24.3<br>(7.9, 74.9) | <.0001 | 23.6<br>(9.4, 59.5) | <.0001 |
|  |  | Cognetta | 4 | 3.0 | 129 | 4.2<br>(1.0, 17.1) | 0.0438 | 4.1<br>(1.2, 14.2) | 0.0259 |
|  |  | All | 33 | 10.8 | 273 | 14.8<br>(5.1, 43.3) | <.0001 | 14.4<br>(6.1, 33.9) | <.0001 |
| SCCIS | US-SRT | Yu (2021) | 2 | 0.5 | 415 | OR | p-value | OR | p-value |
|  |  | Moloney (2022) | 1 | 0.2 | 649 |  |  |  |  |
|  | XRT/SRT | Cognetta | 19 | 2.2 | 842 | 4.7<br>(1.1, 20.2) | 0.0385 | 14.6<br>(2.0, 109.7) | 0.0090 |

Notes: Silverman presented 2-year and 5-year follow-up results. Mean 5-year follow-up was used. Mean follow-up for Yu, Moloney, Lovett, Locke, and Cognetta, respectively, was 2.1, 2.1, 5, 5 and 2.6 years. Silverman presented no SCC data. Only Cognetta presented SCCIS.

Table 1 indicates that US-SRT LC was statistically superior to the comparator XRT/SRT studies individually, collectively, and stratified by histologic tumor type, with p-values ranging from p<0·0001 to p= 0·0438. Figure 1 plots the LC probabilities for each US-SRT study compared to each XRT/SRT study separated by histology. Figure 1 affirms graphically the findings shown in Table 1.

**Figure 1.**
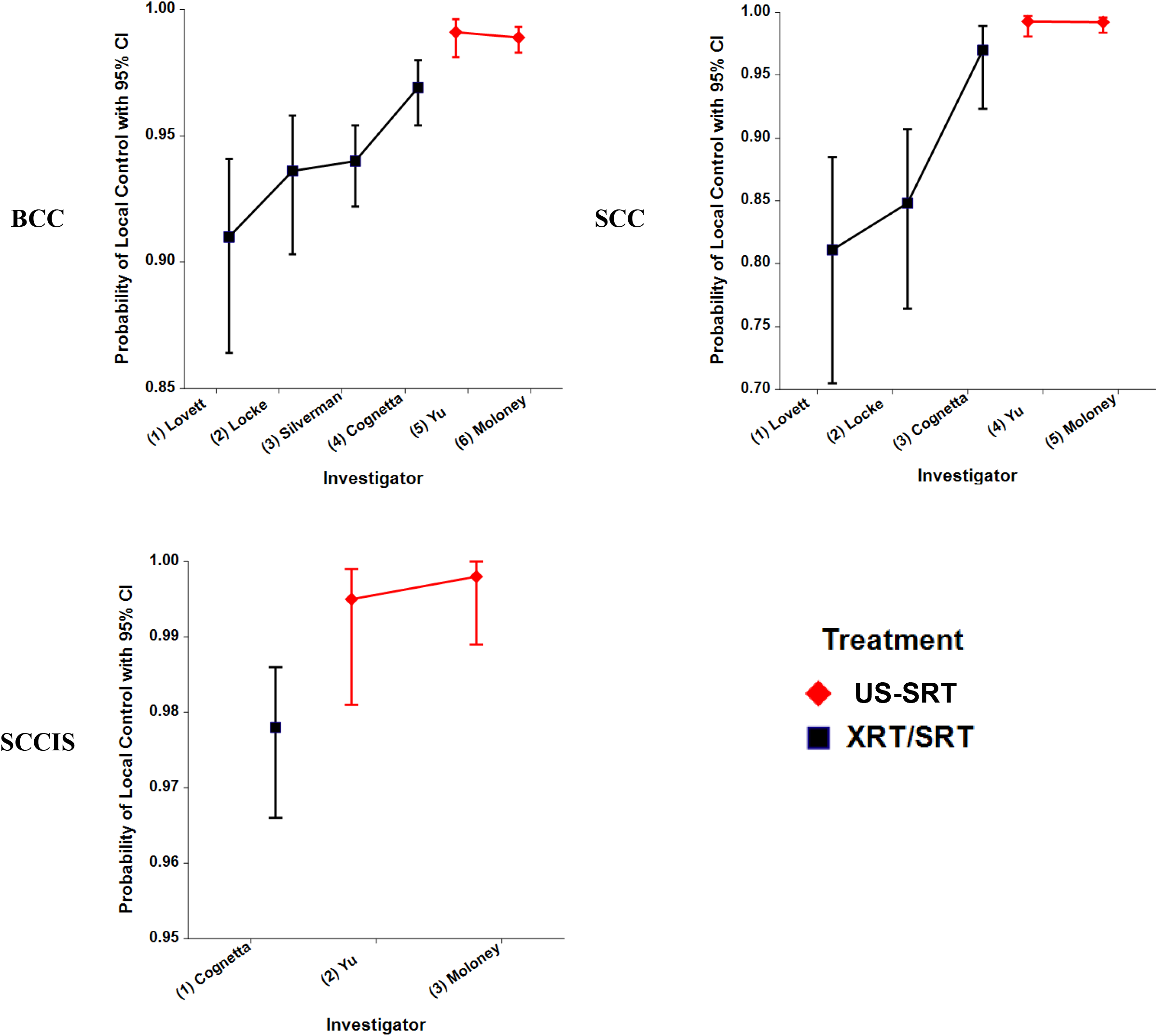
Basal Cell Carcinoma (BCC), Squamous Cell Carcinoma (SCC) and Squamous Cell Carcinoma In Situ (SCCIS) Probabilities of Local Control for the High resolution dermal ultrasound image guided superficial radiotherapy (US-SRT) and External Radiation Therapy (XRT) / Superficial Radiation Therapy (SRT) Investigations

## Discussion

Our analysis shows US-SRT confers a statistically significant improvement in local control for all histologic subtypes of epithelial NMSC (BCC, SCC, SCCIS) compared to all four high-quality, recent, large studies using non-image-guided forms of radiotherapy (XRT/SRT). The improvement in local control can be attributed to the image-guided component of superficial radiotherapy, as the high definition integrated dermal ultrasound with doppler features allows for visualization of the early-stage lesions’ depth, breadth, and overall configuration prior to, during, and after treatment. Visualization during treatment allows for the treatment provider to adjust radiotherapy dosages and energies of penetration daily if necessary. Visualization after treatment allows for confirmation of lesion resolution/response.

Advantages of US-SRT include high cure-rates as demonstrated in the 2021 and 2022 papers. It is cost effective, due to its low-recurrence rate. It offers cosmetic benefits as it is tissue sparing and the majority of NMSC lesions occur in cosmetically sensitive areas, such as the head and neck. Patients can avoid pain, scarring and risk of infection and/or bleeding since this is a non-surgical treatment modality. As many patients often have more than one lesion diagnosed simultaneously, multiple lesions can also be treated synchronously with US-SRT. Offices that use US-SRT have reported overall excellent (> 95%) patient satisfaction and provider satisfaction (internal data).

Absolute and relative contraindications to US-SRT, as previously described in the 2021 US-SRT studies, include lesion invasion to underlying bone or muscle, thickness > 6mm, previous radiation to the same lesion site, ataxia telangiectasia, active connective tissue disease, active lupus or rheumatologic disease, concomitant management with radiation sensitizing chemotherapy agent, T4 stage, and node positive status. [8]

US-SRT could be considered the preferred standard non-surgical radiotherapeutic treatment modality for appropriate patients with early-stage epithelial skin cancers (BCC, SCC, SCCIS) with comparable LC to MMS without the drawbacks of surgery. At the minimum, patients should be presented with the option to have their NMSC treated with US-SRT.

## Limitations

No randomized controlled trial exists for direct comparison of US-SRT to radiotherapy modalities, including superficial and external radiotherapy. The follow-up periods in this paper, though long enough to reasonably assure meaningful and accurate US-SRT to XRT/SRT comparisons, are unequal among studies.

## Conclusion

Image guidance with high resolution dermal ultrasound in the form of US-SRT is shown to confer a statistically significant advantage in lesion local control over non-image guided forms of SRT or XRT in all subtypes of cutaneous epithelial NMSC and should be considered the preferred standard of non-surgical treatment for early stage cutaneous BCC, SCC, and SCCIS.

## Data Availability

All data produced in the present study are available upon reasonable request from the corresponding author, Dr. Lio Yu, at. Data will be available for request with publication for period of at least 1 year. Additional analyses may be available upon request, including but not limited to all odds and probabilities using logistic regression, conversion of some of the odds to probabilities with asymmetric confidence limits, raw percentages, and raw odds.

## List of Abbreviations

SRT: Superficial Radiotherapy
MHz: Megahertz
LC: Local Control
ASTRO: American Society of Radiation Oncology
BCC: Basal Cell Carcinoma
SCC: Squamous Cell Carcinoma
SCCIS: Squamous Cell Carcinoma in-Situ
NMSC: Non-Melanoma Skin Cancer
MMS: Mohs Micrographic Surgery
PDT: Photodynamic Therapy
EBRT: Electron Beam Radiotherapy
XRT: External Radiation Therapy
IGSRT: Image-Guided Superficial Radiation Therapy

## Declarations

### Ethics approval and consent to participate

This study was performed in compliance with the pertinent sections of the Helsinki Declaration and its amendments. All methods were carried out in accordance with relevant guidelines and regulations. The protocol for this manuscript was reviewed by the WCG IRB Affairs Department located at 1019 39^th^ Ave SE, Suite 120, Puyallup, WA 98374 and was determined to be exempt from IRB approval under 45 CFR 46.104 (d)(4), because the research involves the use of identifiable private information; and information is recorded by the investigator in such a manner that the identity of the human subject cannot readily be ascertained directly or through identifiers linked to the subjects, the investigator does not contact the subjects, and the investigator will not re-identify subjects.

Any health information used in this study has been de-identified for use in this study. All patients gave informed consent before treatment.

### Consent for publication

Not applicable

### Availability of data and materials

Deidentified data are available on request from the corresponding author – Dr. Lio Yu at. Data will be available for request with publication for period of at least 1 year.

Additional analyses may be available upon request from Dr. Lio Yu at, including but not limited to all odds and probabilities using logistic regression, conversion of some of the odds to probabilities with asymmetric confidence limits, raw percentages, and raw odds.

### Authors’ contributions

Drafts of the manuscript were shared among the authors. All authors read and approved the final manuscript.

Dr. Lio Yu is responsible for conceptualization of the study design, methodology, data curation, funding acquisition, project administration, writing – original draft, writing – review & editing, supervision, investigation, visualization, and resources. Mairead Moloney was responsible for the literature search, writing – original draft, and writing – review & editing, and visualization. Songzhu Zheng was responsible for formal analysis, validation, and software. Dr. James Rogers was responsible for formal analysis, validation, software, writing – review & editing, figures, investigation, and visualization.

Songzhu Zheng directly accessed and verified the underlying data reported in the manuscript. Dr. James Rogers had access to the underlying data and applied statistical analysis to the data in tabulated format. The logistic analysis was validated under Summit Analytical SOPs.

No medical writer or editor was used or involved in preparing this paper for publication.

## Acknowledgements

–We would like to thank the authors of the studies used for comparison analysis in this manuscript and for data made available for this purpose. We would like to thank the patients for their information used in this study.

## Footnotes

A Per the American Academy of Dermatology practice guidelines, Mohs micrographic survey (MMS) local control is reported to have local control (LC) of 99% for basal cell carcinoma (BCC) and about 97% for cutaneous squamous cell carcinoma (SCC). [4] [6] The 5-year local recurrence rate for primary SCC lesions treated with MMS is reported to be 3.1%. The 5-year local recurrence rate for primary BCC lesions treated with MMS is reported to be 1%. Our reported high resolution dermal ultrasound image guided superficial radiotherapy LC is 99.1% (2021 US-SRT study^8^) and 98.9% (2022 US-SRT study^9^) for BCC and is 99.3% (2021 US-SRT study^8^) and 99.2% (2022 US-SRT study^9^) for SCC, and thus appears to be as good as MMS for BCC and potentially better for SCC.

B Minor differences may be present in the raw numbers of each histologic subtype [BCC, SCC, SCCIS] used in the original analysis by Yu et al. and the raw numbers used in this analysis.^8^ These differences can be attributed to variations in data filtering. For instance, follow-up data was calculated in days and converted to weeks and months, thus data may be filtered by follow-up time in days, weeks, or months. Additionally, seven lesions had multiple histologic subtypes and can be included in both histology categories in this analysis. Despite these minor differences in total lesion number per histologic category, the overall local control rate results remained substantively unchanged.

